# Estimated benefit of PCSK9 inhibition following ischemic stroke due to severe intracranial atherosclerosis

**DOI:** 10.1101/2024.10.31.24316532

**Authors:** James E. Siegler, Elena Badillo Goicoechea, Shadi Yaghi, Rami Z. Morsi, Andrea Arevalo, Matthew M. Smith, Sachin Kothari, Harsh Desai, Neha Sehgal, Rohini Rana, Caroline Kellogg, Aditya Jhaveri, Adam de Havenon, Therese Dunne, Kamil Cameron, Seemant Chaturvedi, Malik Ghannam, Shyam Prabhakaran, Elisheva Coleman, James R. Brorson, Rachel Mehendale, Tareq Kass-Hout

**Author notes:** Corresponding Author: James E. Siegler University of Chicago 5841 S. Maryland Ave MC 2030, S243 Chicago, IL 60637 X: @JimSiegler.

## Abstract

**Background:** The risk of recurrent atherosclerotic vascular events in patients with stroke due to intracranial atherosclerotic disease (ICAD) is high. Proprotein convertase subtilisin/kexin type 9 inhibitors (PCSK9i) can dramatically lower low-density lipoprotein (LDL) levels when added to statins, but are not currently indicated for patients with ICAD.

**Methods:** In this secondary analysis of the SAMMPRIS trial, we estimated the association between LDL reduction (enrollment to day 30, by quartiles) and recurrent cerebral infarction or myocardial infarction (MI) beyond 30 days (primary outcome).

Estimates were assessed using adjusted Cox proportional hazards regression accounting for age, sex, race, Hispanic ethnicity, baseline LDL, vascular comorbidities, and statin use prior to enrollment. We applied relative LDL reduction estimates from PCSK9i trials to project adjusted incidence rate differences of the primary outcome observed in SAMMPRIS with an equivalent LDL reduction. Semiparametric Cox models estimated the annualized relative risk of primary events if PCSK9i were used in the SAMMPRIS population.

**Results:** Of the 451 patients from SAMMPRIS, 378 met inclusion criteria. By day 30, LDL levels fell significantly (absolute change -19mg/dL, IQR -41 to -2). In unadjusted Cox regression compared to the lowest quartile, patients in the highest quartile of LDL improvement trended toward a lower rate of the primary outcome (Q4 vs. Q1 hazard ratio [HR] 0.66, 95% confidence interval [CI] 0.35-1.26), achieving significance after multivariable adjustment (adjusted HR 0.38, 95% CI, 0.16-0.89). Every 10 mg/dL improvement in 30-day LDL was associated with a 9% lower rate of the primary outcome (adjusted HR 0.91, 95% CI, 0.83-0.997). Assuming an average projected effect of PCSK9i, with half of SAMMPRIS patients having been treated, PCSK9i use could reduce the annualized risk of the primary outcome by 33.2%.

**Conclusions:** More aggressive LDL lowering in patients with stroke due to ICAD is associated with lower rates of recurrent stroke or MI, and this can be achieved with PCSK9i use.

## BACKGROUND

The effect of high-intensity statins in preventing recurrent cerebrovascular events among patients with stroke due to atherosclerosis is largely driven by the individual responsiveness to statin therapy and reduction in low-density lipoprotein cholesterol (LDL). Lower LDL levels in patients with and without prior cerebral infarction have strongly and independently correlated with a lower risk of atherosclerotic vascular events. One prior meta-analysis reported a reduction of ∼5% of such events for every ∼10 mg/dL lower LDL.^1^ High-intensity statin medications such as atorvastatin and rosuvastatin can significantly reduce serum LDL levels in patients,^2^ with atorvastatin reducing LDL by a median of 1.57 mmol/L (95% CI, 1.31-2.07), and rosuvastatin reducing LDL by a median of 1.74 mmol/L (95% CI, 0.83-2.50) versus placebo.^3^

When used in conjunction with high-intensity statins, novel lipid-lowering therapies such as proprotein convertase subtilisin/kexin type 9 inhibitors (PCSK9i) can lead to a greater decrease in LDL levels. Pooled data from the ODYSSEY trials (Evaluation of Cardiovascular Outcomes After an Acute Coronary Syndrome During Treatment With Alirocumab) reported a mean 55% relative reduction in LDL, or a mean absolute difference of 66 mg/dL with alirocumab when added to maximally tolerated, high-intensity statin.^4^ Similar findings were observed in the FOURIER trial with the addition of evolocumab when added to statin, with an associated reduction in cardiovascular events.^5^ Moreover, PCSK9i have been shown to reduce coronary atheroma volume, which has not been observed with high-intensity statin treatment.^6^ Based on these data, the Food and Drug Administration has approved alirocumab and evolocumab for adults with clinical atherosclerotic disease who require additional lowering of LDL levels.

In this post-hoc analysis of the Stenting and Aggressive Medical Management for Preventing Recurrent Stroke in Intracranial Stenosis (SAMMPRIS) trial, we evaluated vascular event risk according to improvement in LDL levels at follow-up. From these relationships, we extrapolated the potential benefit of PCSK9i use in a high-risk population with atherosclerosis.

## METHODS

The SAMMPRIS trial protocol was approved by the institutional review boards of participating sites, US Food and Drug Administration, and Data and Safety Monitoring Board appointed by the NIH. Informed consent was obtained from each participant prior to enrollment. Data from the SAMMPRIS trial can be made available to a qualified investigator upon reasonable request of the National Institute of Neurological Disorders and Stroke. The results of this analysis are reported in accordance with the Strengthening the Reporting of Observational Studies in Epidemiology Guidelines.^7^

### Patient population

Please refer to the original results of SAMMPRIS for detailed information regarding patient enrollment and inclusion.^8^ Briefly, SAMMPRIS was an investigator-initiated randomized clinical trial conducted across 50 sites in the United States. Patients were eligible for inclusion if they experienced an acute ischemic stroke or transient ischemic attack related to intracranial atherosclerosis with 70-99% luminal stenosis in an artery proximal to the territory of ischemia. In SAMMPRIS, patients were randomized 1:1 to undergo intracranial arterial stenting with dual antiplatelet therapy (aspirin and clopidogrel) for 90 days, or to receive aspirin and clopidogrel for 90 days. All patients were managed aggressively for underlying vascular risk factors, including treatment with a high-intensity statin (rosuvastatin) to a target LDL of <70 mg/dL, antihypertensive agent(s) to achieve a goal systolic blood pressure of <140 mmHg (<130 mmHg for patients with diabetes), and additional diet and lifestyle modifications as described in the trial protocol.^9^ Patients underwent formal evaluations on day 4, day 30, and every 4 months after enrollment unless a primary endpoint would occur, in which follow-up would occur 90 days thereafter. Patients underwent repeat laboratory testing in the central laboratory for lipid levels at day 30 and 4 months, with repeat testing thereafter made at the discretion of the investigator based on recent LDL level.^9^

In this secondary analysis, patients were eligible for inclusion if they were treated without intracranial stenting and if they had at least one follow-up measurement of LDL and lipoprotein(a) levels, as well as complete covariates for the multivariable modeling described below. Given the primary exposure variable is the change in LDL level from baseline to initial follow-up (approximately 30 days after enrollment), patients with a primary outcome event occurring within 30 days were excluded from this analysis.

### Covariates

The primary exposure of interest was the absolute change in LDL from enrollment to day 30, divided according to quartiles. Comparisons were primarily made between the uppermost quartile of LDL improvement versus the lowermost quartile due to possible non-linear association of the exposure. Secondary exposures included absolute LDL reduction as a continuous variable (in 1mg/dL and 10mg/dL increments), relative change in LDL by day 30 (as a percent change from baseline), and absolute and relative changes in LDL from baseline to 4 months for those with 4-month laboratory results. Additional covariates included age at enrollment, sex, statin use within 7 days of enrollment, baseline lipid levels, baseline hemoglobin A1c, history of hypertension, diabetes, myocardial infarction (MI), congestive heart failure, stroke, and active tobacco use (defined as use within 6 months of enrollment).

### Outcomes

The primary outcome was a composite of recurrent cerebral infarction or MI. Cerebral infarction was defined as a symptomatic or asymptomatic cerebral infarction according to the data dictionary provided by the trial investigators. There was no standardized definition for asymptomatic infarction provided in the trial protocol.^8^

### Statistical analyses

Descriptive statistics were used to summarize patient characteristics and changes in laboratory parameters from baseline evaluation to first follow-up.

A Cox proportional hazards model was fitted to estimate the risk of the primary outcome according to quartiles of LDL improvement from baseline to first follow-up. This model was subsequently adjusted for age, sex, body mass index, race, Hispanic ethnicity, hypertension, diabetes, MI, congestive heart failure, statin use at enrollment, tobacco use, baseline National Institutes of Health Stroke Scale (NIHSS) score, and baseline LDL level. In a sensitivity analysis, we repeated the adjusted Cox proportional hazards regression including quartile of LDL reduction from baseline to 30d (and separately, baseline to 4 months) as the primary independent covariate. The exposure effect (LDL reduction) on the primary outcome was assessed across the following pre-specified subgroups: intracranial arterial stenting versus no stenting, age (by tertile), sex, prior statin use versus no statin use, and baseline LDL level (by tertile). Heterogeneity of exposure effect was assessed across these subgroups using a multiplicative interaction term for absolute LDL reduction (30d versus baseline) x covariate of interest in the multivariable Cox model for the primary outcome. The proportional hazards assumption was evaluated visually using log-log plots and confirmed via testing the scaled Shoenfeld residuals.

The theoretical benefit of PCSK9i was assessed by comparing adjusted incidence rates of the primary outcome in the SAMMPRIS population according to actual versus projected LDL reduction scenarios, where projected LDL reduction varied according to: 1) baseline LDL levels, 2) different proportions of patients from SAMMPRIS assumed to be prescribed PCSK9i, and 3) hypothetical PCSK9i administration assuming baseline treatment effect of PCSK9i on LDL consistent with recent evidence. To achieve this, we first fitted a Poisson regression for the primary outcome using the aforementioned demographic and clinical covariates, plus relative change in LDL from baseline to day 30 as the primary exposure. The expected change in LDL was in turn projected as described previously, incorporating the average effect of PCSK9i treatment on LDL reduction (∼60%) from a network meta-analysis of 48 randomized clinical trials^10^ into the same fitted model. The estimated incidence rate for the primary outcome was then evaluated for this threshold of relative LDL change using predictive margins. Additional thresholds were also tested to estimate mild and robust theoretical treatment effects (relative changes in LDL of -40% and -80%). Estimated differences in annualized incidence rates for all nine scenarios were thus calculated across the included cohort by contrasting the projected change in primary outcomes under PCSK9i versus no PCSK9i (i.e. using the original LDL changes observed in the SAMMPRIS data) for each individual in the sample, all other factors left constant in the model. To account for the potential diminishing marginal benefits of PCSK9i on LDL (e.g., for patients starting with already low LDL levels, further PCSK9i-induced reductions would be plausibly very modest), the projected effect was further made to be logarithmically dependent on each patient’s LDL baseline level. Calculations were repeated presuming the effects were observed for 25% of the cohort, and for 50% of the cohort, presuming only these proportions of patients from SAMMPRIS would have been treated with PCSK9i.

Analyses were performed using STATA v18.0 (College Station, TX) and R v4.3.2. All tests were performed at the two-sided level with an alpha of 0.05. No adjustments were made for multiple hypothesis testing.

## RESULTS

Of the 451 patients from SAMMPRIS, 378 patients had complete covariate data for this secondary analysis and were included (Figure 1). Compared to included patients, those who were excluded were older (median 64 vs. 59y, p<0.01), more frequently female (57.5% vs. 36.2%, p<0.01), and had more concomitant diabetes (54.8% vs. 40.2%, p=0.02; See Online Supplement). Among included patients, the median baseline LDL was 91 mg/dl (IQR 72-116), with a majority (86%) of patients taking a statin at the time of enrollment. Across quartiles of change in LDL from baseline to day 30, patients with a greater improvement (reduction) in LDL had higher baseline LDL levels and were less frequently taking a statin prior to trial enrollment (Table 1).

**Figure 1.**
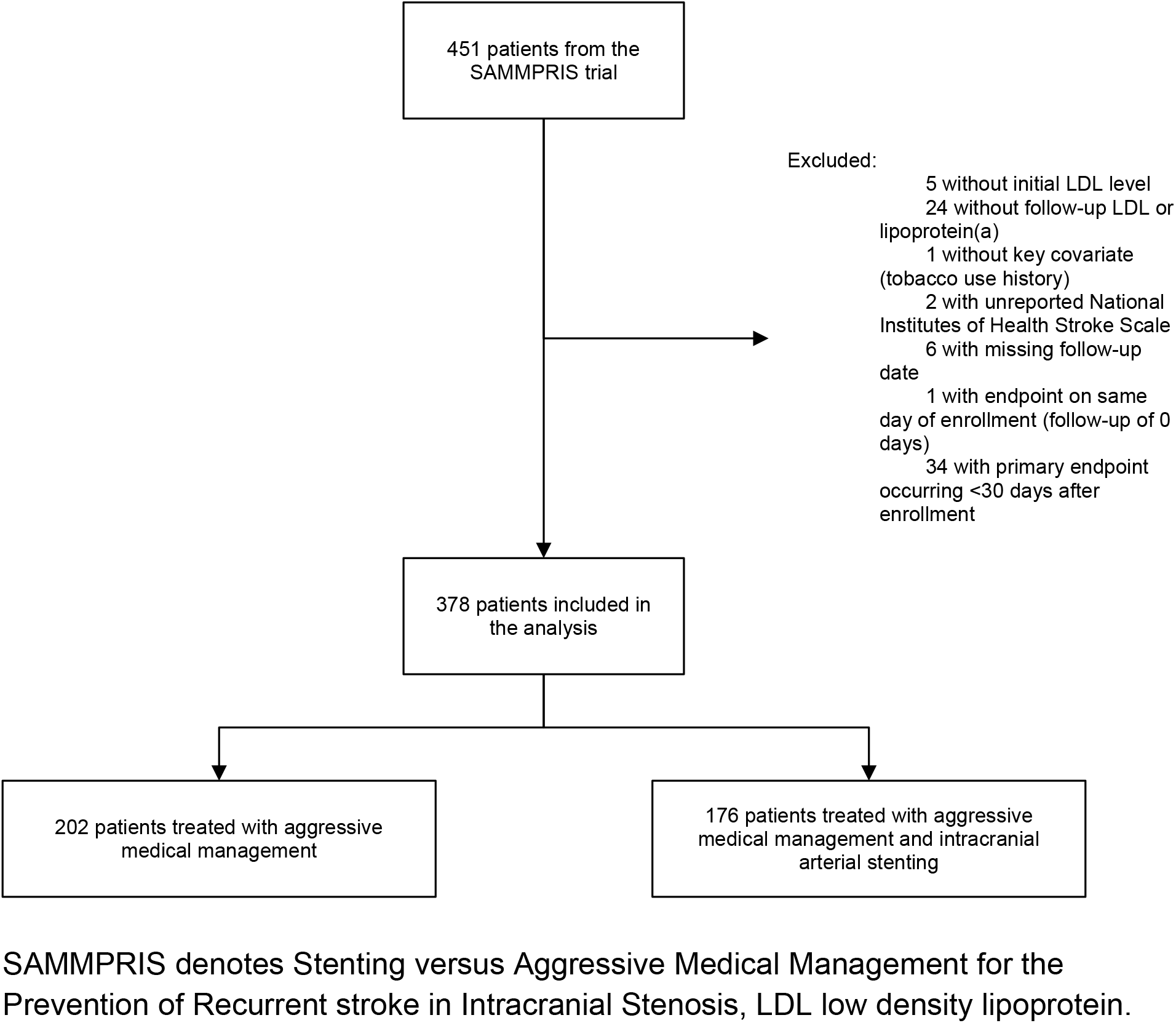
Inclusion flowchart.

**Table 1.**
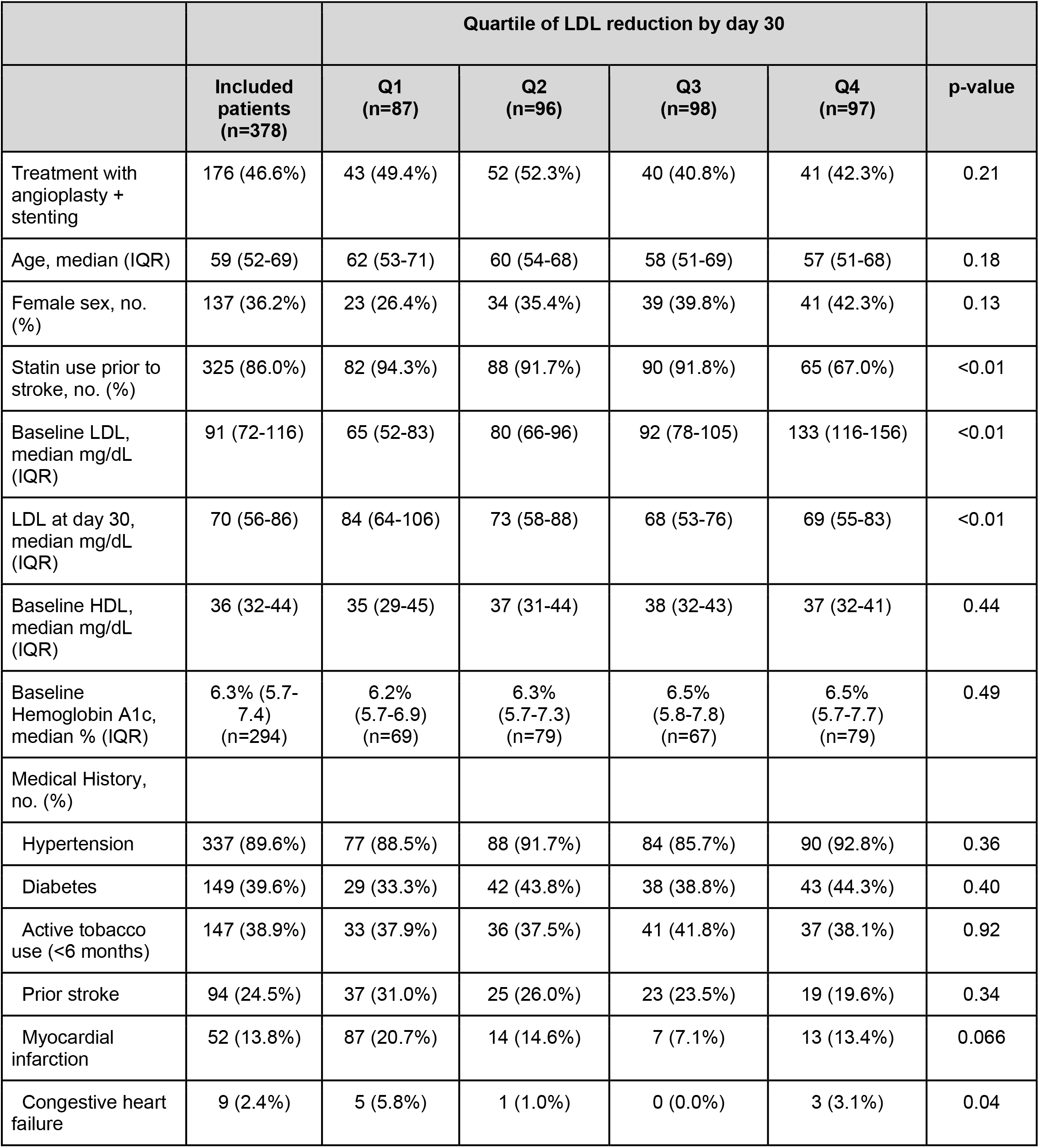
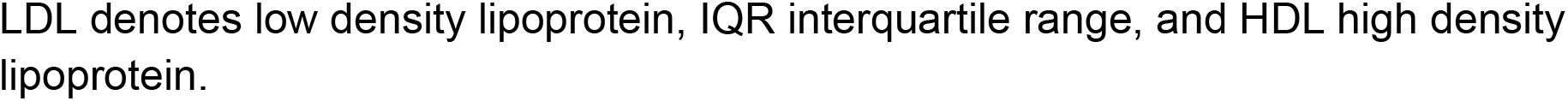
Patient characteristics stratified by quartile of LDL reduction.

Across the cohort, the majority of patients experienced an improvement in LDL by day 30 (n=278, 75.9%). Two hundred forty-seven patients (65.3%) achieved an LDL reduction of at least 10% by 30 days. The median absolute reduction in LDL by day 30 was 19 mg/dL for the cohort (IQR -41 to -2; Table 2) with a similar change in LDL sustained by 4 months among those with repeat testing (n=356/378 of the included cohort, median change -20 mg/dL, IQR -44 to -4).

**Table 2.** Change in LDL from baseline.

|  |  | Quartile of LDL reduction |  |  |  |  |
| --- | --- | --- | --- | --- | --- | --- |
|  | Included patients<br>(n=378) | Q1<br>(n=87) | Q2<br>(n=96) | Q3<br>(n=98) | Q4<br>(n=97) | p-value |
| Relative change in LDL by day 30, % (IQR) | -22.4%<br>(-38.0 to -2.0) | +16.4%<br>(6.8 to 41.1) | -13.3%<br>(-18.1 to -5.1) | -29.0%<br>(-33.7 to -26.7) | -47.8%<br>(-57.6 to -31.4) | <0.01 |
| Absolute change in LDL by day 30, mg/dL (IQR) | -19<br>(-41 to -2) | +10<br>(+4 to +30) | -10<br>(-14 to -4) | -26<br>(-32 to -23) | -62<br>(-80 to -49) | <0.01 |
| Relative change in LDL by 4 months, % (IQR) | -25.6%<br>(-42.2 to -5.7)<br>(n=356) | -0.6%<br>(-16.5 to +20.7) | -17.1%<br>(-27.8 to -5.6) | -27.3%<br>(-36.7 to -10.7) | -46.6%<br>(-59.4 to -36.2) | <0.01 |
| Absolute change in LDL by 4 months, mg/dL (IQR) | -20<br>(-44 to -4)<br>(n=356) | -0.5<br>(-10 to +13) | -14<br>(-22 to -5) | -24.5<br>(-37 to -11) | -59.5<br>(-83 to -44) | <0.01 |

LDL denotes low density lipoprotein and IQR interquartile range.

The total analysis time was 972.8 patient-years, during which a primary endpoint occurred for 67 patients (17.7%) over a median follow-up of 236 days (IQR 131-506). The incidence rate of the primary outcome was 68.9 per 1,000 patient-years (95% CI 54.2-87.5). In unadjusted Cox proportional hazards regression, there was a non-significant reduction in rate of the primary outcome for quartiles 2-4 when compared to the lowermost quartile of LDL improvement (compared to Q1, Q2 hazard ratio [HR] 0.55, 95% confidence interval [CI] 0.28-1.08; Q3 HR 0.58, 95% CI 0.30-1.13; Q4 HR 0.66, 95% CI, 0.35-1.26). Following multivariable adjustment, the difference between Q4 and Q1 became statistically significant (adjusted HR [aHR] 0.38, 95% CI, 0.16-0.89, p=0.03; Figure 2). In that model, statin use prior to enrollment was not independently associated with the primary outcome (aHR 0.60, 95% CI, 0.30-1.18).

**Figure 2.**
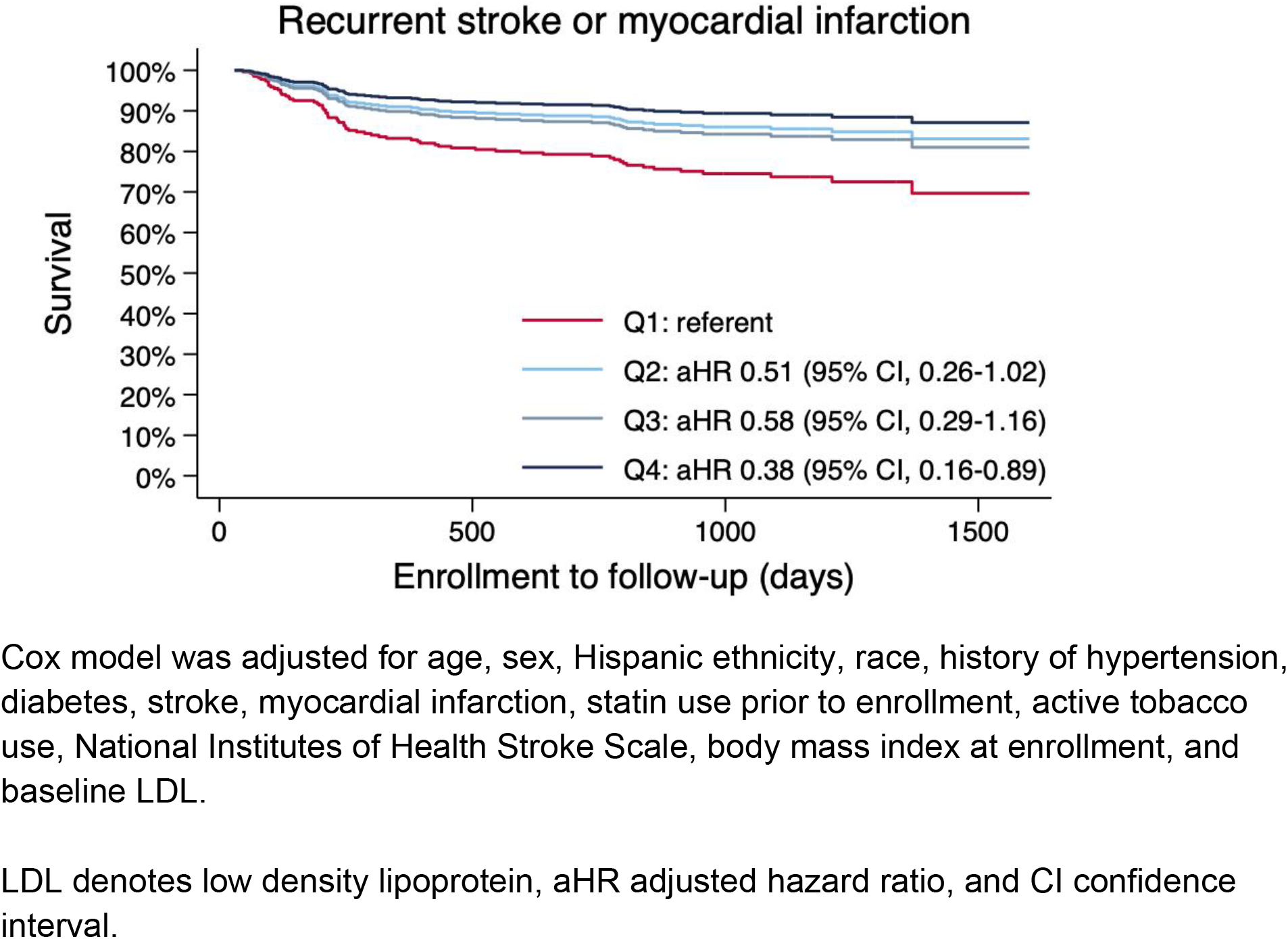
Survival curve for the primary outcome according to absolute LDL reduction by day 30.

In unadjusted Cox regression, considering LDL change as a continuous exposure, every 10 mg/dL improvement in LDL at 30 days was not significantly associated with a reduction in the rate of the primary outcome (HR 0.98, 95% CI, 0.91-1.05, p=0.62).

Following multivariable adjustment, every 10 mg/dL LDL improvement (reduction) was independently associated with a 9% lower rate of the primary outcome (aHR 0.91, 95% CI, 0.83-0.997, p=0.043). There were no statistically significant interactions between LDL lowering and pre-specified subgroups for the primary outcome (Table 3). Even for patients taking statin prior to enrollment, there was a significant beneficial effect with additional LDL lowering (aHR for every 10mg/dL LDL reduction: 0.89, 95% CI, 0.80- 0.99).

**Table 3.** Subgroup analysis of treatment effect for every 10mg/dL LDL reduction by day 30.

|  | Adjusted HR (95% CI) | p-value | p<sub>interaction</sub> |
| --- | --- | --- | --- |
| Trial intervention |  |  | 0.13 |
| Best medical management | 0.83 (0.73-0.95) | <0.01 |  |
| Angioplasty + stenting | 0.96 (0.81-1.12) | 0.58 |  |
| Age (tertile of years) |  |  | 0.27 |
| Tertile 1 (32-55) | 0.92 (0.78-1.09) | 0.35 |  |
| Tertile 2 (55-66) | 0.96 (0.79-1.16) | 0.66 |  |
| Tertile 3 (66-81) | 0.81 (0.67-0.97) | 0.02 |  |
| Sex |  |  | 0.09 |
| Male | 0.91 (0.80-1.03) | 0.15 |  |
| Female | 0.86 (0.73-1.02) | 0.08 |  |
| Statin prior to enrollment |  |  | 0.12 |
| No | 1.08 (0.83-1.40) | 0.58 |  |
| Yes | 0.89 (0.80-0.99) | 0.03 |  |
| Tertile of baseline LDL (mg/dL) |  |  | 0.19 |
| Tertile 1 (31-78) | 0.89 (0.72-1.10) | 0.29 |  |
| Tertile 2 (79-107) | 0.80 (0.63-1.00) | 0.052 |  |
| Tertile 3 (108-261) | 0.95 (0.84-1.08) | 0.43 |  |

Hazard ratios calculated for the primary outcome of recurrent cerebral infarction or myocardial infarction.
LDL denotes low density lipoprotein, aHR adjusted hazard ratio.

Assuming modest, average, and high projected effects of PCSK9i (40%, 60%, and 80% reduction in LDL), with half of SAMMPRIS patients having been treated, PCSK9i use could have reduced the annualized risk of the primary endpoint by 23.2%, 33.2%, and 41.2%, respectively (Table 4). Assuming all SAMMPRIS patients were treated and an average PCSK9i effect, the primary outcome would have been reduced by 51.9%.

**Table 4.** Estimates of relative risk reduction of primary endpoint with PCSK9i.

|  |  | Projected LDL lowering effect based on meta-analyzed trials comparing PCSK9 inhibitors versus placebo among statin-users |  |  |
| --- | --- | --- | --- | --- |
|  |  | Modest projected LDL lowering effect (40% relative reduction) | Average projected LDL lowering effect (60% relative reduction)<sup>10</sup> | High projected LDL lowering effect (80% relative reduction) |
| Theoretical proportion of patients from SAMMPRIS prescribed PCSK9i | 10% | -3.29% | -5.76% | -7.78% |
|  | 25% | -6.47% | -12.83% | -17.99% |
|  | 50% | -23.18% | -33.24% | -41.23% |
|  | 75% | -30.19% | -43.40% | -53.51% |
|  | 100% | -36.50% | -50.89% | -61.58% |
|  |  | Cells represent projected annualized incidence rate change in the expected primary endpoint with PCSK9i administration versus no PCSK9i administration, every other observed baseline characteristic held constant. |  |  |

PCSK9i denotes proprotein convertase subtilisin/kexin type 9 inhibitor, LDL low density lipoprotein, and SAMMPRIS Stenting versus Aggressive Medical Management for Preventing Recurrent stroke in Intracranial Stenosis.

## DISCUSSION

Consistent with prior data, we observed a dose-response relationship between LDL lowering and the risk of recurrent ischemic stroke or MI among patients treated with high-intensity statin. Notably, patients from the SAMMPRIS trial had generally low LDL levels at baseline, likely owing to the fact that a majority were already prescribed a statin, as compared to other observational cohorts with stroke due to intracranial atherosclerosis.^11^ It is possible that the lower baseline LDL levels in SAMMPRIS patients may have attenuated the effect of LDL reduction in this population. Even so, a significant association was observed between larger LDL improvements and a reduction in stroke or MI risk. When stratified by quartiles of LDL improvement, all patients in the first quartile experienced an increase in LDL by 30 days. Compared to patients in this quartile, those in every other quartile of LDL improvement had a lower risk of the primary outcome, with patients in the uppermost quartile showing the most significant risk reduction.

The magnitude of the association between lower LDL levels and the primary outcome is worth emphasizing. Risk factor control in the medical arm of SAMMPRIS has already been reported to strongly predict subsequent vascular events.^12^ In this analysis of the larger SAMMPRIS cohort, the annualized incidence rate of the primary outcome of nearly 70 per 1,000 patients (while high) is lower than would be expected for this population due to the exclusion of patients with events occurring within the first 30 days. In our analysis, we excluded 34 patients with a primary endpoint occurring during this hyperacute window, at which point the theoretical effect of LDL reduction and potential benefit of PCSK9i would be negligible. These additional 34 patient events would increase the event rate by 50%. Even so, the lower LDL reduction during the initial 30 days after the incident stroke was associated with a dramatic reduction in recurrent atherosclerotic events. Even a 10% reduction in LDL from baseline–which was achieved in two-thirds of the cohort–was associated with a 10% reduction in the primary outcome.

In addition to the magnitude of the association, the timing of aggressive LDL lowering in relation to the primary outcome warrants further explanation. In this study, we found no significant additional LDL lowering between 30 and 120 days after enrollment. LDL lowering within the first month was strongly associated with early and sustained reduction in the primary outcome event. We do not believe this relationship is due to differences in patient treatment in SAMMPRIS, as all participants received the highest tolerated dose of a high-intensity statin, along with regular counseling from diet and lifestyle coaches. We cannot know for certain if there were differences in compliance with statin and diet and/or lifestyle recommendations between each quartile of LDL reduction in this analysis. However, it is possible that nonadherence to one or more interventions played a role. Lower LDL levels to the degree observed in SAMMPRIS were likely driven largely by treatment with statins, which are known to have pleiotropic effects on the vascular system.^13^ In addition to the inhibition of cholesterol biosynthesis in the liver, statins can reduce systemic inflammation (as biomarkers such as c-reactive protein^14^ and reduced leukocyte trafficking and T-cell migration^15^), may improve endothelial function and possess antiplatelet properties.^1613^ Each of these mechanisms may occur more rapidly than LDL reduction (which may only be a “biomarker” of early statin effects on the endothelium) and may mediate the favorable outcomes observed in this analysis.

It is likely that the majority of the LDL reduction, and the differential response (variable LDL changes), was driven by the use and tolerance of high-intensity statin therapy. LDL improvement may represent statin use and response, and therefore LDL changes could be a surrogate indicator of the pleiotropic response to statin treatment. Subsequently, any reduction in stroke risk may be indirectly linked to non-LDL lowering effects of statins rather than the observed LDL improvements observed at the patient level.

Unfortunately, adherence to and dosing of the statin agent at follow-up visits are not available, so a mediation analysis of LDL lowering in response to statin cannot be conducted. That said, the majority of patients in SAMMPRIS were already taking a statin medication at the time of trial enrollment, although significantly fewer patients in the uppermost quartile of LDL reduction at 30 days were taking statins. In this subgroup, there remained a statistically significant reduction in the primary outcome with lower LDL levels (Table 3). Furthermore, given the presumably high rate of statin use and tolerance throughout the trial period, the pleiotropic effects of statins would be similarly distributed throughout the cohort.

As shown in Figure 2, there is a significant reduction in the risk of recurrent cerebral infarction or MI with even modest reductions in LDL. Although the difference across quartiles was non-significant in unadjusted regression, the effect estimates were in the direction of a beneficial effect with greater LDL reductions. Because this effect became significant after multivariable adjustment, we believe this is largely due to residual confounding. There were meaningful differences between patient groups according to LDL reduction quartiles, including lower proportion of statin treatment prior to enrollment among Q4 versus Q1. While there was no significant interaction observed between statin use at enrollment and LDL reduction for the primary outcome, it is possible that the greater reduction LDL levels and in primary outcome events was related to an increase in statin use in this patient group. That said, statin pretreatment was not independently associated with the outcome in the multivariable model.

The more dramatic lowering of LDL with PCSK9i has the potential to lead to major reductions in event rates for patients with stroke due to ICAD. Using data from published clinical trials demonstrating an average 60% relative reduction in LDL levels with PCSK9i use, we estimated the additional, theoretical benefit of PCSK9i if used concomitantly with high-intensity statin treatment and other aggressive medical management in SAMMPRIS. With PCSK9i use, the absolute risk of recurrent stroke or MI events >30 days after incident stroke due to intracranial atherosclerosis could be halved.^17^ Large artery intracranial atherosclerotic disease accounts for 10-15% of ischemic strokes worldwide.^18^ In the United States, we estimate this may include as many as 80-100,000 patients per year. If even a modest reduction in LDL is achieved for a minority of patients treated with PCSK9i for intracranial atherosclerotic disease, there may be enormous clinical benefits. These are measurable at the patient level, with a significant individual reduction of atherosclerotic vascular events, hospital utilization and costs of care, resultant disability, and ultimately improved survival. Moreover, the beneficial effects of PCKS9i may exceed the LDL-lowering effect. These agents are known to lower lipoprotein(a) levels,^19^ and may have local antiplatelet and anti-inflammatory effects at sites of plaque formation.^20^

Among the pre-specified subgroups tested in this analysis, we observed no significant interaction between absolute LDL reduction and the primary outcome. Therefore, we are left to conclude that there is no difference in potential benefit of greater LDL reduction for preventing stroke or MI across age, sex, prior statin use, baseline LDL level, or use of intracranial stenting. That said, greater LDL reductions in the subgroup of patients treated with best medical management (without intracranial stenting) were associated with a lower risk of the primary outcome. By contrast, LDL reductions among stented patients resulted in a nearly neutral effect estimate. It is likely that the heightened risk of in-stent thrombosis despite dual antiplatelet therapy may have attenuated any treatment benefit of LDL reduction, plaque stabilization, or other effect of statin and diet/lifestyle modifications.

### Limitations

The greatest limitation to this analysis is the small sample size. That said, a significant association between LDL reduction and lower outcome events was observed, largely owing to the high risk condition of intracranial atherosclerosis. As stated previously, the patient characteristics among participants in SAMMPRIS are not perfectly reflective of what may be seen in clinical practice. Many patients already had reasonable LDL levels at the time of enrollment, with approximately half of patients having an LDL <90 mg/dL. In our subgroup analysis according to baseline LDL level, we did not observe a significant interaction between change in LDL and the primary outcome across tertiles of baseline LDL. Although an interaction was not observed in our small sample size, it is possible that a larger cohort of patients (with more patients having higher LDL levels) may observe greater benefit with aggressive LDL lowering. The effect of LDL reduction in cardiovascular events can be appreciated in trials conducted over several years, however the effect may not be as significant in the early window. In SAMMPRIS, recurrent stroke events were measured at 3 and 12 months, which may have been too early to appreciate an effect of LDL reduction with statin use and other secondary prevention recommendations.

### Conclusions

In this secondary analysis of the SAMMPRIS trial, in which there was regular assessment of LDL levels and standardized lipid-lowering interventions among patients with high-risk atherosclerotic vascular disease, we observed improved outcomes with LDL lowering by 30 days. Additional LDL lowering, even beginning from reasonable or low baseline levels, is associated with a lower risk of recurrent stroke or MI. The addition of novel lipid-lowering therapies, such as PCSK9i and other interventions, to maximally tolerated statin therapy and diet/lifestyle counseling has the potential to further and dramatically reduce the risk of atherosclerotic vascular events. In high-risk patients, such as those with stroke due to intracranial atherosclerotic disease, the use of such adjuvant therapies may be warranted, even when currently accepted LDL targets are reached.

## Data Availability

Data from the SAMMPRIS trial will be made available by the NINDS upon reasonable request.

## ACKNOWLEDGMENTS

The authors would like to thank the NINDS for making these data available for this analysis.

